## Supplement Tables and Figures for "Performance of Multimodal GPT-4V on USMLE with Image: Potential for Imaging Diagnostic Support with Explanations"

### Supplementary Figure 1. A case on how GPT-4V answers medical license questions with images. We provide the link to question, GPT-4V response, and analysis provided by healthcare professional.

Link to question: <https://next.amboss.com/us/questions/qjBb0CYawe/8>

AMBOSS step1 Qa3

GPT -4V Response (Prompt without Hint)

The chest X-ray provided appears to show a large area of translucency (darkness) on the patient's right side, suggesting the presence of air outside the lung but inside the chest cavity. This is consistent with a pneumothorax, in which air accumulates between the lung and the chest wall, causing the lung to collapse.

Given the clinical presentation of sudden onset shortness of breath in a young woman who doesn't have other risk factors (e.g., smoking, recreational drug use), this could be a spontaneous pneumothorax. Tall, thin individuals are at higher risk for spontaneous primary pneumothorax.

Given this, let's interpret the potential physical examination findings:

**A) Absent vesicular sounds, dull percussion, asymmetric chest expansion, decreased tactile fremitus**:

This combination doesn't match the X-ray findings. With a pneumothorax, we'd expect absent or decreased breath sounds, but not dullness on percussion.

**B) Absent breath sounds, hyperresonant percussion, asymmetric chest expansion, decreased tactile fremitus**:

This is consistent with a pneumothorax. The affected side would show absent breath sounds because the collapsed lung is not ventilating. Percussion would be hyperresonant due to the presence of air. The affected side would have decreased chest expansion and decreased tactile fremitus because vibration transmission would be affected by the pneumothorax.

**C) Fine inspiratory crackles, dull percussion, symmetric chest expansion, increased tactile fremitus**:

This is more consistent with pneumonia or pulmonary edema, not a pneumothorax.

**D) Bronchial breath sounds, dull percussion, asymmetric chest expansion, increased tactile fremitus**:

Bronchial breath sounds and increased tactile fremitus might be seen in consolidation, such as in pneumonia, but not with a pneumothorax.

**E) Decreased vesicular sounds, hyperresonant percussion, symmetric chest expansion, decreased tactile fremitus**:

This option presents with hyperresonance similar to a pneumothorax, but the symmetric chest expansion is inconsistent with the X-ray and clinical presentation.

The correct set of findings for this patient, given the X-ray and clinical scenario, is:

**B) Absent breath sounds, hyperresonant percussion, asymmetric chest expansion, decreased tactile fremitus**.

GPT -4V Response (Prompt with Hint)
The chest x-ray findings, along with the clinical context provided, are suggestive of a severe asthma exacerbation. Let's correlate the expected physical examination findings in an asthma attack with the given options:

1. **Chest auscultation**: During an asthma attack, bronchoconstriction leads to decreased air entry and often wheezing. In severe attacks, airflow may be so reduced that breath sounds become very diminished or even absent.

2. **Chest percussion**: Due to the trapped air in the lungs, percussion would be hyperresonant.

3. **Chest expansion**: Asthma attacks can cause asymmetric chest expansion if one part of the lung is more affected than the other. However, symmetric decreased chest expansion can also be seen when both lungs are equally affected.

4. **Tactile fremitus**: Fremitus is decreased when there's more air (as in hyperinflation) between the surface and lung parenchyma.

Given these findings, the best matching option is:

**E. Decreased vesicular sounds, hyperresonant, symmetric, decreased tactile fremitus**.

Let's dissect each option and understand the clinical scenarios or conditions that could lead to these sets of findings:

**A. Absent vesicular sounds, dull percussion, asymmetric chest expansion, decreased tactile fremitus**

- **Absent vesicular sounds**: No air movement, which can be seen in a complete obstruction or collapse (atelectasis) of the lung.

- **Dull percussion**: Atelectasis, pleural effusion, or pneumonia, which lead to consolidation or fluid build-up.

- **Asymmetric chest expansion**: One lung or part of a lung is not expanding as well as the other side, possibly due to unilateral diseases like pleural effusion, pneumothorax, or atelectasis.

- **Decreased tactile fremitus**: Sound waves don't transmit well through fluid or air, hence conditions like pleural effusion or pneumothorax can lead to decreased fremitus.

**B. Absent breath sounds, hyperresonant percussion, asymmetric chest expansion, decreased tactile fremitus**

- **Absent breath sounds**: Can be due to severe obstruction or total collapse of the lung.

- **Hyperresonant percussion**: Presence of excessive air, as in pneumothorax.

- **Asymmetric chest expansion**: One lung is affected more than the other, as seen in a unilateral pneumothorax.

- **Decreased tactile fremitus**: Air does not transmit vibrations well, as in pneumothorax.

**C. Fine inspiratory crackles, dull percussion, symmetric chest expansion, increased tactile fremitus**

- **Fine inspiratory crackles**: Usually seen in conditions like early congestive heart failure or interstitial lung diseases.

- **Dull percussion**: Suggests consolidation or fluid, like in pneumonia.

- **Symmetric chest expansion**: Both lungs expand equally.

- **Increased tactile fremitus**: Sound waves travel better through solid media. Pneumonia, which is a consolidation of the lung, can lead to increased fremitus.

**D. Bronchial breath sounds, dull percussion, asymmetric chest expansion, increased tactile fremitus**

- **Bronchial breath sounds**: Normally heard over the trachea but when heard over lung fields, it suggests underlying consolidation, as in pneumonia.

- **Dull percussion**: Again, indicative of consolidation or fluid.

- **Asymmetric chest expansion**: One side affected more than the other, possibly due to unilateral pneumonia.

- **Increased tactile fremitus**: Consolidation from pneumonia allows for better transmission of sound waves.

**E. Decreased vesicular sounds, hyperresonant percussion, symmetric chest expansion, decreased tactile fremitus**

- **Decreased vesicular sounds**: Diminished but not absent, indicative of conditions like asthma or early COPD where airflow is decreased but not completely absent.

- **Hyperresonant percussion**: Air trapping as seen in asthma or COPD.

- **Symmetric chest expansion**: Both lungs are equally affected, as often seen in asthma or COPD.

- **Decreased tactile fremitus**: The hyperinflated lungs with increased air do not transmit the vibrations well.

In the context of the presented case, option E is consistent with the findings seen in an asthma exacerbation.

Expert Analysis Below:

This case demonstrates an error response on a hard question with chest x-ray, as patient symptoms in the question stem matched with both option B pneumothorax and option E asthma. GPT-4V misunderstood the X-ray by showing pneumothorax because of the translucency on the right lung which indicates air outside the lung but inside the chest cavity and thus gave the wrong answer. Then we provide GPT-4V with the hint that interprets this image: *The chest x-rays show increased radiolucency, flattened diaphragm, and wide intercostal spaces, all of which are signs of pulmonary hyperinflation.* GPT-4V corrected its answer to option E, which is the correct answer from AMBOSS.

### Supplementary Figure 2. A case on how GPT-4V interacts with physicians. We provide the physician question and GPT-4V response.

**Question:**

A 45-year-old woman was admitted to this hospital because of dyspnea on exertion, fatigue, and confusion. The patient had been in her usual state of health until 18 months before the current admission, when the blood pressure was noted to be 140/90 mm Hg during a routine appointment with her primary care physician. Therapy with oral contraceptive pills was discontinued, but the blood pressure was persistently elevated when it was measured at home and during subsequent clinical evaluations. Results of duplex ultrasonography of the bilateral renal arterial vasculature were normal, as were levels of thyrotropin, plasma free catecholamines, plasma metanephrines, and aldosterone. Transthoracic echocardiography revealed normal ventricular function, with no evidence of valvular disease or left ventricular hypertrophy; the interventricular septal wall thickness was 11 mm (normal range, 7 to 11). Lisinopril was administered, and the blood pressure decreased to 120/80 mm Hg.

Four weeks before the current admission, swelling of the legs, abdominal distention, intermittent dyspnea on exertion, and fatigue developed, and the patient was evaluated in the emergency department of this hospital. She reported a weight gain of 4.5 kg during the previous month, occasional episodes of confusion, and two episodes of burning chest pain during exercise that had lasted for 2 minutes each and were relieved with rest. On examination, the temperature was 36.8°C, the blood pressure 180/110 mm Hg, the heart rate 73 beats per minute, the respiratory rate 18 breaths per minute, and the oxygen saturation 98% while the patient was breathing ambient air. The weight was 66.9 kg; approximately 5 months earlier, at a clinic visit, it had been 67.1 kg. The body-mass index (the weight in kilograms divided by the square of the height in meters) was 22. A systolic murmur (grade 2/6) was best heard at the left sternal border. The jugular venous pressure was estimated to be 10 cm of water. The abdomen was soft, nontender, and slightly distended. There was trace nonpitting edema in the legs. The remainder of the physical examination was normal. An electrocardiogram showed a normal sinus rhythm and left-axis deviation. Laboratory test results are shown in Table 1. A chest radiograph showed a normal cardiac silhouette and small bilateral pleural effusions, with no opacities or pulmonary edema. The patient was admitted to this hospital. The patient had a history of endometriosis and dysmenorrhea, conditions that had been treated with a levonorgestrel-releasing intrauterine system. She had a remote history of palpitations that were not associated with evidence of arrhythmia on Holter monitoring. The only medication was lisinopril. The patient’s mother and sister had both had venous thromboses and miscarriages. Her maternal uncle had hypertrophic cardiomyopathy; there was no family history of sudden cardiac death. The patient did not drink alcohol, smoke tobacco, or use illicit drugs. She was married and worked as a lawyer.

Furosemide was administered intravenously, and the patient reported a decrease in the dyspnea and swelling. Daily therapy with a low dose of oral furosemide was initiated. A transthoracic echocardiogram showed hyperdynamic left ventricular systolic function, with an ejection fraction of 76% (normal range, 50 to 75), as well as biventricular hypertrophy, an interventricular septal wall thickness of 14 mm, mild mitral regurgitation, systolic anterior motion of the mitralvalve leaflets, and accelerated flow in the left ventricular outflow tract. Because the blood pressure was persistently elevated, the dose of lisinopril was increased and treatment with metoprolol and nifedipine was initiated. On the third hospital day, the patient was discharged home.

Three weeks before the current admission, dyspnea on exertion and fatigue recurred, and the patient was again evaluated in the emergency department of this hospital. She reported a 3-day history of malaise and night sweats. On examination, the temperature was 36.8°C, the blood pressure 148/96 mm Hg, the heart rate 84 beats per minute, the respiratory rate 16 breaths per minute, and the oxygen saturation 96% while she was breathing ambient air. The weight was 65.3 kg. Jugular venous pulsation was not visible, and there was no leg edema; otherwise, the results of the physical examination had not changed from the previous week. Laboratory test results are shown in Table 1. Blood and urine cultures were sterile. Tests for human immunodeficiency virus (HIV) type 1 p24 antigen and HIV type 1 and type 2 antibodies were negative. An interferon-γ release assay for tuberculosis was nonreactive. A chest radiograph showed that the pleural effusions had resolved and that no opacities or pulmonary edema had developed. The patient was admitted to this hospital, and additional cardiac imaging studies were obtained.

CT was performed after the administration of intravenous contrast material. In a few sentences, what disease does the sagittal image (Panel C) indicate?

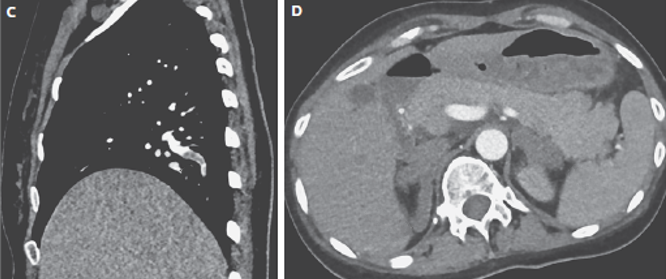

**GPT response**:

The sagittal image (Panel C) appears to show a computed tomography (CT) scan of the chest. The image suggests the presence of a ground-glass opacification, which is a radiological term that describes an area of increased attenuation in the lung without obscuration of the underlying vascular markings. Ground-glass opacities can be seen in a variety of conditions, but in the context of recent clinical presentations and findings, it is often associated with viral pneumonias, including COVID-19.

**Question**:

In a few sentences, what disease does the arrow in sagittal image (Panel C) indicate?

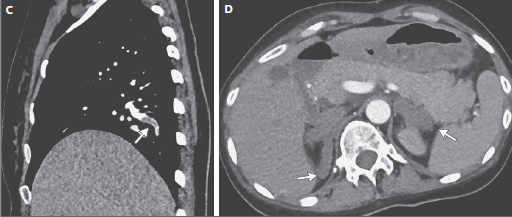

**GPT response**:

The arrow in the image might support pulmonary embolism (PE). A PE occurs when a blood clot, usually from the legs or pelvis (known as deep vein thrombosis or DVT), travels to the lungs and obstructs one of the pulmonary arteries or its branches. When reviewing a CT image, the presence of a filling defect within the contrast-enhanced pulmonary artery can indicate a PE. The segmental artery in the right lower lobe, as indicated by the arrow, seems to have such a defect, making it suggestive of a PE.

**Question**: In a few sentences, what disease does the arrow in the axial image (Panel D) indicate?

**GPT response:**

The arrow in the axial image (Panel D) points to an abnormality in the adrenal gland. Given the clinical presentation and the findings on the CT scan, the abnormality suggests a diagnosis of an adrenal adenoma, which can lead to primary aldosteronism or Conn's syndrome. This condition is characterized by excessive aldosterone secretion by the adrenal glands, which can result in hypertension and electrolyte imbalances.

**Question**:

The patient’s prescribed dose of oral furosemide was administered, but orthostatic hypotension developed. She received intravenous fluids, and furosemide therapy was discontinued. She did not have a fever during the hospitalization. The white-cell count decreased but did not normalize. On the sixth hospital day, the patient was discharged home. The weight was 62.1 kg. At home, the fatigue continued to decrease until 2 weeks after discharge, when recurrent fatigue, fevers, and palpitations developed. The patient was again evaluated in the emergency department of this hospital. The patient’s husband reported that she had become increasingly confused, irritable, and anxious. He also noted that she appeared to be weak globally, with increased respiratory effort on exertion. The patient reported frequent urination and increased thirst. On examination, the temperature was 36.2°C, the blood pressure 135/80 mm Hg, the heart rate 98 beats per minute, the respiratory rate 18 breaths per minute, and the oxygen saturation 98% while the patient was breathing ambient air. The weight was 60.0 kg. The patient was awake, alert, and cooperative, but she was unable to state the year or to count backward from 100 in increments of 7. She occasionally responded to questions inappropriately and with confabulation. Cranial-nerve function was normal, although the first cranial nerve was not tested. Strength was 4/5 in the proximal muscles of the arms and legs. Sensation was normal. A systolic murmur (grade 3/6) was best heard at the left sternal border. Jugular venous pulsation was not visible, and there was no leg edema. The amount of scalp hair had decreased, and hair growth on the upper lip and chin had increased. She had no rashes, bruises, striae, or hyperpigmentation. The remainder of the physical examination was normal. Laboratory test results are shown in the Table.

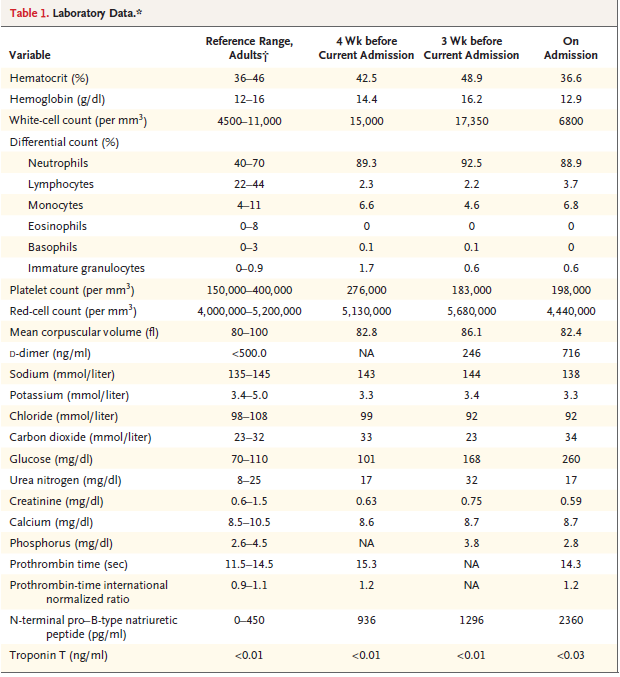

What are the differential diagnoses on the possible causes of severe hypertension for this patient? Please list out 6 most possible differential diagnoses and your rationale, including the patient symptoms and lab results to support your diagnoses.

**GPT response**:

Based on the provided patient history, symptoms, physical examination, and laboratory data, here are six possible differential diagnoses for the cause of hypertension in this patient, along with rationales:

1. **Primary Aldosteronism (Conn's syndrome)**

**Rationale**: The patient's hypertension despite the discontinuation of oral contraceptive pills, along with her normal bilateral renal arterial vasculature and normal levels of plasma free catecholamines, metanephrines, and aldosterone, may hint towards this. The CT finding in the adrenal gland further supports this possibility.

2. **Cushing's Syndrome**

**Rationale**: The patient exhibits symptoms such as increased hair growth on the upper lip and chin, decreased scalp hair, no hyperpigmentation, and a potential adrenal abnormality on CT. The lab results indicate elevated glucose levels, which can be a manifestation of Cushing's syndrome.

3. **Pheochromocytoma**

**Rationale**: The palpitations, anxiety, and high blood pressure might indicate this, but the normal catecholamine levels make it less likely. The adrenal abnormality seen on CT can also be due to pheochromocytoma.

4. **Thyroid Dysfunction (particularly hyperthyroidism or thyrotoxicosis)**

**Rationale**: Symptoms such as palpitations, fatigue, weight loss, increased hair growth, and mental changes can be associated with thyroid dysfunction. Additionally, the patient’s increased heart rate and the noted systolic murmur at the left sternal border could be suggestive of hyperthyroidism.

5. **Renovascular Hypertension**

**Rationale**: While the duplex ultrasonography of the bilateral renal arterial vasculature was reported as normal, it's still a consideration given the severe hypertension.

6. **Polycystic Ovary Syndrome (PCOS)**

**Rationale**: The increased hair growth on the upper lip and chin, along with decreased scalp hair, could be indicative of hirsutism, which is often seen in PCOS. However, other classic signs of PCOS are not mentioned.

It's important to note that these are potential diagnoses based on the provided information, and additional testing and evaluation are essential to pinpoint the exact cause of the patient's hypertension.

**Question**:

If I suspect Cushing’s syndrome due to ectopic corticotropin secretion, what is the next step in this patient’s evaluation to identify the source of abnormal hormone production?

**GPT response**:

If you suspect Cushing's syndrome due to ectopic corticotropin (ACTH) secretion, the next steps in the patient’s evaluation to identify the source of abnormal hormone production are:

1. **Measurement of ACTH Levels**:

Before determining the source of ectopic ACTH production, you should first confirm that the Cushing's syndrome is ACTH-dependent. This is done by measuring serum ACTH levels. Elevated ACTH levels would suggest ACTH-dependent Cushing's syndrome, which can be due to either Cushing's disease (pituitary origin) or ectopic ACTH secretion (non-pituitary origin).

2. **High-Dose Dexamethasone Suppression Test**:

In this test, a high dose of dexamethasone is given, and then cortisol levels are measured. In Cushing's disease (pituitary source of excess ACTH), cortisol levels typically suppress with high-dose dexamethasone. In ectopic ACTH secretion, cortisol levels usually do not suppress.

3. **Cranial Imaging**:

If ACTH levels are elevated, perform an MRI of the pituitary gland to rule out pituitary adenomas causing Cushing's disease. If no pituitary adenoma is detected, it increases the suspicion for ectopic ACTH secretion.

4. **Ectopic Source Localization**:

If ectopic ACTH secretion is suspected, the next step is to localize the source of the ectopic production. Potential sources include bronchial carcinoids, small cell lung cancer, medullary thyroid cancer, and other neuroendocrine tumors.

**Chest CT or MRI**: To check for lung tumors, especially small cell lung cancer or carcinoid tumors.

**Abdominal and Pelvic CT or MRI**: To check for tumors in the abdomen or pelvis that could be secreting ACTH.

**Octreotide or Gallium-Dotatate PET/CT Scans**: These scans can be useful to identify neuroendocrine tumors, which might be the source of ectopic ACTH.

5. **Inferior Petrosal Sinus Sampling (IPSS)**:

If there's still uncertainty after the above steps, IPSS can differentiate between ectopic ACTH secretion and Cushing's disease. In this procedure, blood samples are taken from the veins that drain the pituitary gland and are compared to blood samples from a peripheral vein. A significantly higher ACTH level in the petrosal sinuses compared to the peripheral blood would suggest a pituitary source.

After the source of ectopic ACTH production is identified, appropriate management, which might include surgical resection of the tumor, can be initiated.

**Question**:

Axial PET-CT images of the lower chest obtained before and after the administration of contrast material (Panels A and B of the image, respectively). Is there support to verify the previous suspicion?

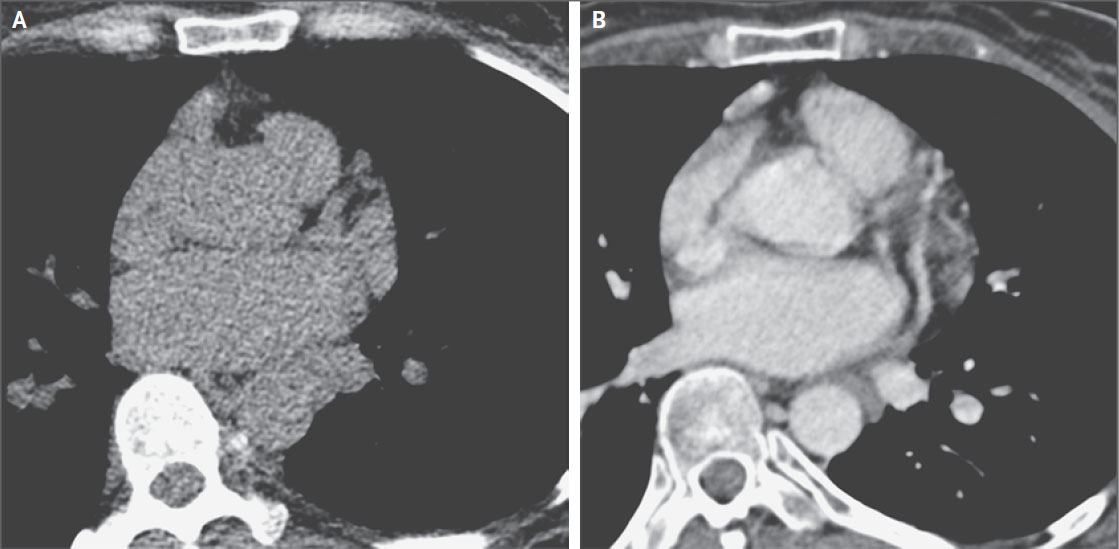

**GPT response**:

The images you've provided are axial PET-CT scans of the lower chest. Based on the images:

- **Panel A** shows an unenhanced (or pre-contrast) scan. The main feature that stands out is a large mass or lesion in the mediastinal region. The mass appears heterogeneous with some areas of increased density, possibly suggesting a mix of solid and necrotic/cystic components.

- **Panel B** shows the post-contrast image of the same region. The lesion demonstrates enhancement after the administration of contrast material, suggesting the presence of blood vessels within the lesion. The enhancement pattern might help in characterizing the lesion further.

Given the context you provided regarding a suspicion of ectopic corticotropin (ACTH) secretion (potentially due to a tumor secreting ACTH), the presence of a mediastinal mass could be significant. The mediastinum is a known site for neuroendocrine tumors like bronchial carcinoids, which can sometimes lead to ectopic ACTH secretion and subsequent Cushing's syndrome.

These images seem to provide support to the suspicion of a mediastinal source for ectopic ACTH secretion. However, a definitive diagnosis would require further studies, such as a biopsy of the mass, to confirm its nature and to determine if it's the source of ectopic ACTH secretion. Always consult with a radiologist and an endocrinologist for a comprehensive evaluation.

### Supplementary Table 1. Performance of GPT-4V across different subdomains.

| **URL** | **GPT4v Reponse** | **Correct Response** | **Discipline** | **Status Without hint** |
| --- | --- | --- | --- | --- |
| <https://next.amboss.com/us/exam-simulation/MWdb0MNKse/1> | D | D | Family Medicine | Correct |
| [https://next.amboss.com/us/questions/iVdb0J8Kse/1](https://next.amboss.com/us/exam-simulation/MWdb0MNKse/2) | C | E | Family Medicine | Incorrect |
| <https://next.amboss.com/us/questions/iVdb0J8Kse/2> | D | H | Family Medicine | Incorrect |
| <https://next.amboss.com/us/questions/iVdb0J8Kse/3> | A | A | Family Medicine | Correct |
| <https://next.amboss.com/us/questions/iVdb0J8Kse/4> | D | D | Family Medicine | Correct |
| <https://next.amboss.com/us/review/MWdb0MNKse/3> | B | B | Family Medicine | Correct |
| <https://next.amboss.com/us/questions/pTdb0L76se/1> | E | E | Neurology | Correct |
| <https://next.amboss.com/us/questions/pTdb0L76se/2> | D | C | Neurology | Incorrect |
| <https://next.amboss.com/us/questions/pTdb0L76se/3> | D | C | Neurology | Incorrect |
| <https://next.amboss.com/us/questions/LRdb0wnpse/1> | C | D | Pharmacology and Toxicology | Incorrect |
| <https://next.amboss.com/us/questions/LRdb0wnpse/2> | A | A | Pharmacology and Toxicology | Correct |
| <https://next.amboss.com/us/questions/LRdb0wnpse/3> | C | E | Pharmacology and Toxicology | Incorrect |
| <https://next.amboss.com/us/questions/Hodb0KWIse/1> | D | C | Emergency Medicine | Incorrect |
| <https://next.amboss.com/us/questions/Hodb0KWIse/2> | C | A | Emergency Medicine | Incorrect |
| <https://next.amboss.com/us/questions/Hodb0KWIse/3> | E | D | Emergency Medicine | Incorrect |
| <https://next.amboss.com/us/questions/Hodb0KWIse/4> | A | A | Emergency Medicine | Correct |
| <https://next.amboss.com/us/questions/Uqdb0bBIse/1> | A | A | Immunology(difficulty 3) | Correct |
| <https://next.amboss.com/us/questions/Uqdb0bBIse/2> | G | G | Immunology(difficulty 3) | Correct |
| <https://next.amboss.com/us/questions/Uqdb0bBIse/3> | D | D | Immunology(difficulty 3) | Correct |
| <https://next.amboss.com/us/questions/Uqdb0bBIse/4> | C | C | Immunology(difficulty 3) | Correct |
| <https://next.amboss.com/us/questions/Uqdb0bBIse/5> | D | D | Immunology(difficulty 3) | Correct |
| <https://next.amboss.com/us/questions/bHdb0HKrse/1> | D | D | Gynecology(Difficulty 2/3) | Correct |
| <https://next.amboss.com/us/questions/bHdb0HKrse/2> | D | D | Gynecology(Difficulty 2/3) | Correct |
| <https://next.amboss.com/us/questions/bHdb0HKrse/3> | D | D | Gynecology(Difficulty 2/3) | Correct |
| <https://next.amboss.com/us/questions/GUVb0BeFse/1> | A | E | Clinical Pharmacology | Incorrect |
| <https://next.amboss.com/us/questions/GUVb0BeFse/2> | A | F | Clinical Pharmacology | Incorrect |
| <https://next.amboss.com/us/questions/GUVb0BeFse/3> | D | D | Clinical Pharmacology | Correct |
| <https://next.amboss.com/us/questions/GUVb0BeFse/4> | D | G | Clinical Pharmacology | Incorrect |
| <https://next.amboss.com/us/questions/GUVb0BeFse/5> | C | C | Clinical Pharmacology | Correct |
| <https://next.amboss.com/us/questions/GUVb0BeFse/6> | C | C | Clinical Pharmacology | Correct |
| <https://next.amboss.com/us/questions/GUVb0BeFse/8> | B | A | Clinical Pharmacology | Incorrect |
| <https://next.amboss.com/us/questions/6SVb0j08se/1> | B | C | Anatomy | Incorrect |
| <https://next.amboss.com/us/questions/6SVb0j08se/2> | D | C | Anatomy | Incorrect |
| <https://next.amboss.com/us/questions/6SVb0j08se/3> | B | E | Anatomy | Incorrect |
| <https://next.amboss.com/us/questions/6SVb0j08se/4> | A | A | Anatomy | Correct |
| <https://next.amboss.com/us/questions/xsVb0ECDse/1> | B | B | Abdominal Surgery | Correct |
| <https://next.amboss.com/us/questions/xsVb0ECDse/2> | F | F | Abdominal Surgery | Correct |
| <https://next.amboss.com/us/questions/xsVb0ECDse/3> | A | A | Abdominal Surgery | Correct |
| <https://next.amboss.com/us/questions/xsVb0ECDse/4> | E | E | Abdominal Surgery | Correct |
| <https://next.amboss.com/us/questions/xsVb0ECDse/5> | E | E | Abdominal Surgery | Correct |
| <https://next.amboss.com/us/questions/xsVb0ECDse/6> | G | C | Abdominal Surgery | Incorrect |
| <https://next.amboss.com/us/questions/xsVb0ECDse/7> | E | E | Abdominal Surgery | Correct |
| <https://next.amboss.com/us/questions/xsVb0ECDse/8> | A | E | Abdominal Surgery | Incorrect |
| <https://next.amboss.com/us/questions/X8Vb09Owse/1> | E | E | Orthopedics | Correct |
| <https://next.amboss.com/us/questions/X8Vb09Owse/2> | F | F | Orthopedics | Correct |
| <https://next.amboss.com/us/questions/X8Vb09Owse/3> | D | D | Orthopedics | Correct |
| <https://next.amboss.com/us/questions/X8Vb09Owse/4> | A | A | Orthopedics | Correct |
| <https://next.amboss.com/us/questions/X8Vb09Owse/5> | E | A | Orthopedics | Incorrect |
| <https://next.amboss.com/us/questions/X8Vb09Owse/6> | A | A | Orthopedics | Correct |
| <https://next.amboss.com/us/questions/X8Vb09Owse/7> | C | C | Orthopedics | Correct |
| <https://next.amboss.com/us/questions/Eieb08tAse/1> | A | A | Rheumatology | Correct |
| <https://next.amboss.com/us/questions/Eieb08tAse/2> | E | G | Rheumatology | Incorrect |
| <https://next.amboss.com/us/questions/Eieb08tAse/3> | A | F | Rheumatology | Incorrect |
| <https://next.amboss.com/us/questions/Eieb08tAse/4> | E | C | Rheumatology | Incorrect |
| <https://next.amboss.com/us/questions/Eieb08tAse/5> | G | G | Rheumatology | Correct |
| <https://next.amboss.com/us/questions/Eieb08tAse/6> | E | E | Rheumatology | Correct |
| <https://next.amboss.com/us/questions/Eieb08tAse/7> | B | D | Rheumatology | Incorrect |
| <https://next.amboss.com/us/questions/OKeb0Ig-se/1> | C | D | Pathology | Incorrect |
| <https://next.amboss.com/us/questions/OKeb0Ig-se/2> | F | A | Pathology | Incorrect |
| <https://next.amboss.com/us/questions/OKeb0Ig-se/3> | A | D | Pathology | Incorrect |
| <https://next.amboss.com/us/questions/OKeb0Ig-se/4> | B | B | Pathology | Correct |
| <https://next.amboss.com/us/questions/OKeb0Ig-se/5> | A | C | Pathology | Incorrect |
| <https://next.amboss.com/us/questions/OKeb0Ig-se/6> | E | E | Pathology | Correct |
| <https://next.amboss.com/us/questions/OKeb0Ig-se/7> | B | C | Pathology | Incorrect |
| <https://next.amboss.com/us/questions/OKeb0Ig-se/8> | D | D | Pathology | Correct |
| <https://next.amboss.com/us/questions/OKeb0Ig-se/9> | D | D | Pathology | Correct |
| <https://next.amboss.com/us/questions/OKeb0Ig-se/10> | D | D | Pathology | Correct |
| <https://next.amboss.com/us/questions/ISUb0Ya1Ge/1> | D | D | Neurosurgery | Correct |
| <https://next.amboss.com/us/questions/ISUb0Ya1Ge/2> | G | F | Neurosurgery | Incorrect |
| <https://next.amboss.com/us/questions/ISUb0Ya1Ge/3> | D | B | Neurosurgery | Incorrect |
| <https://next.amboss.com/us/questions/ISUb0Ya1Ge/4> | B | B | Neurosurgery | Correct |
| <https://next.amboss.com/us/questions/ISUb0Ya1Ge/5> | E | E | Neurosurgery | Correct |
| <https://next.amboss.com/us/questions/ISUb0Ya1Ge/6> | A | A | Neurosurgery | Correct |
| <https://next.amboss.com/us/questions/ISUb0Ya1Ge/7> | B | E | Neurosurgery | Incorrect |
| <https://next.amboss.com/us/questions/QRUb0u51Ge/1> | C | A | behavioural Sciences | Incorrect |
| <https://next.amboss.com/us/questions/ke2b0m_gGe/1> | C | C | Otolaryngology | Correct |
| <https://next.amboss.com/us/questions/ke2b0m_gGe/2> | A | A | Otolaryngology | Correct |
| <https://next.amboss.com/us/questions/ke2b0m_gGe/3> | E | E | Otolaryngology | Correct |
| <https://next.amboss.com/us/questions/ke2b0m_gGe/4> | E | E | Otolaryngology | Correct |
| <https://next.amboss.com/us/questions/ke2b0m_gGe/5> | E | E | Otolaryngology | Correct |
| <https://next.amboss.com/us/questions/ke2b0m_gGe/6> | F | F | Otolaryngology | Correct |
| <https://next.amboss.com/us/questions/Jf2b0sMSGe/1> | A | A | Pulmonology | Correct |
| <https://next.amboss.com/us/questions/Jf2b0sMSGe/2> | D | B | Pulmonology | Incorrect |
| <https://next.amboss.com/us/questions/Jf2b0sMSGe/3> | B | B | Pulmonology | Correct |
| <https://next.amboss.com/us/questions/Jf2b0sMSGe/4> | C | C | Pulmonology | Correct |
| <https://next.amboss.com/us/questions/Jf2b0sMSGe/5> | C | C | Pulmonology | Correct |
| <https://next.amboss.com/us/questions/Jf2b0sMSGe/6> | A | A | Pulmonology | Correct |
| <https://next.amboss.com/us/questions/Jf2b0sMSGe/8> | B | A | Pulmonology | Incorrect |
| <https://next.amboss.com/us/questions/Jf2b0sMSGe/9> | D | D | Pulmonology | Correct |
| <https://next.amboss.com/us/questions/nh2b07ehGe/1> | D | B | Epidemiology and Biostatistics | Incorrect |
| <https://next.amboss.com/us/questions/nh2b07ehGe/2> | D | D | Epidemiology and Biostatistics | Correct |
| <https://next.amboss.com/us/questions/nh2b07ehGe/3> | C | E | Epidemiology and Biostatistics | Incorrect |
| <https://next.amboss.com/us/questions/nh2b07ehGe/4> | C | C | Epidemiology and Biostatistics | Correct |
| <https://next.amboss.com/us/questions/nh2b07ehGe/5> | C | D | Epidemiology and Biostatistics | Incorrect |
| <https://next.amboss.com/us/questions/nh2b07ehGe/6> | B | B | Epidemiology and Biostatistics | Correct |
| <https://next.amboss.com/us/questions/nh2b07ehGe/7> | C | B | Epidemiology and Biostatistics | Incorrect |
| <https://next.amboss.com/us/questions/VQ2b0GEhGe/1> | B | B | Ophthalmology | Correct |
| <https://next.amboss.com/us/questions/VQ2b0GEhGe/2> | D | D | Ophthalmology | Correct |
| <https://next.amboss.com/us/questions/VQ2b0GEhGe/3> | C | C | Ophthalmology | Correct |
| <https://next.amboss.com/us/questions/VQ2b0GEhGe/4> | E | C | Ophthalmology | Incorrect |
| <https://next.amboss.com/us/questions/VQ2b0GEhGe/5> | H | H | Ophthalmology | Correct |
| <https://next.amboss.com/us/questions/VQ2b0GEhGe/6> | E | E | Ophthalmology | Correct |
| <https://next.amboss.com/us/questions/sk2b0t63Ge/1> | E | F | Occupational Medicine | Incorrect |
| <https://next.amboss.com/us/questions/sk2b0t63Ge/2> | B | B | Occupational Medicine | Correct |
| <https://next.amboss.com/us/questions/sk2b0t63Ge/3> | D | D | Occupational Medicine | Correct |
| <https://next.amboss.com/us/questions/sk2b0t63Ge/4> | F | F | Occupational Medicine | Correct |
| <https://next.amboss.com/us/questions/sk2b0t63Ge/5> | E | E | Occupational Medicine | Correct |
| <https://next.amboss.com/us/questions/6l2b0jB3Ge/1> | B | B | Cardiology and angiology | Correct |
| <https://next.amboss.com/us/questions/6l2b0jB3Ge/2> | B | A | Cardiology and angiology | Incorrect |
| <https://next.amboss.com/us/questions/6l2b0jB3Ge/3> | E | E | Cardiology and angiology | Correct |
| <https://next.amboss.com/us/questions/6l2b0jB3Ge/4> | B | B | Cardiology and angiology | Correct |
| <https://next.amboss.com/us/questions/6l2b0jB3Ge/5> | A | F | Cardiology and angiology | Incorrect |
| <https://next.amboss.com/us/questions/6l2b0jB3Ge/6> | B | D | Cardiology and angiology | Incorrect |
| <https://next.amboss.com/us/questions/6l2b0jB3Ge/7> | E | E | Cardiology and angiology | Correct |
| <https://next.amboss.com/us/questions/6l2b0jB3Ge/8> | F | A | Cardiology and angiology | Incorrect |
| <https://next.amboss.com/us/questions/K52b0UORGe/1> | C | D | Critical Care | Incorrect |
| <https://next.amboss.com/us/questions/K52b0UORGe/2> | F | B | Critical Care | Incorrect |
| <https://next.amboss.com/us/questions/K52b0UORGe/3> | C | C | Critical Care | Correct |
| <https://next.amboss.com/us/questions/K52b0UORGe/4> | G | G | Critical Care | Correct |
| <https://next.amboss.com/us/questions/K52b0UORGe/5> | F | B | Critical Care | Incorrect |
| <https://next.amboss.com/us/questions/cn2b0aHRGe/1> | G | C | General Surgery | Incorrect |
| <https://next.amboss.com/us/questions/cn2b0aHRGe/2> | H | H | General Surgery | Correct |
| <https://next.amboss.com/us/questions/cn2b0aHRGe/3> | F | E | General Surgery | Incorrect |
| <https://next.amboss.com/us/questions/cn2b0aHRGe/4> | E | E | General Surgery | Correct |
| <https://next.amboss.com/us/questions/cn2b0aHRGe/5> | C | C | General Surgery | Correct |
| <https://next.amboss.com/us/questions/cn2b0aHRGe/6> | E | E | General Surgery | Correct |
| <https://next.amboss.com/us/questions/cn2b0aHRGe/7> | E | D | General Surgery | Incorrect |
| <https://next.amboss.com/us/questions/cn2b0aHRGe/8> | A | A | General Surgery | Correct |
| <https://next.amboss.com/us/questions/cn2b0aHRGe/9> | B | B | General Surgery | Correct |
| <https://next.amboss.com/us/questions/k62b0mOiGe/1> | E | B | Geriatrics | Incorrect |
| <https://next.amboss.com/us/questions/k62b0mOiGe/2> | D | A | Geriatrics | Incorrect |
| <https://next.amboss.com/us/questions/k62b0mOiGe/3> | D | D | Geriatrics | Correct |
| <https://next.amboss.com/us/questions/k62b0mOiGe/4> | B | B | Geriatrics | Correct |
| <https://next.amboss.com/us/questions/k62b0mOiGe/5> | B | B | Geriatrics | Correct |
| <https://next.amboss.com/us/questions/Nx2b0-9PGe/1> | B | C | Aging | Incorrect |
| <https://next.amboss.com/us/questions/Nx2b0-9PGe/2> | B | E | Aging | Incorrect |
| <https://next.amboss.com/us/questions/4B2b03a4Ge/1> | E | G | Pain Medicine | Incorrect |
| <https://next.amboss.com/us/questions/7B2b04X4Ge/1> | C | E | Infectious Diseases | Incorrect |
| <https://next.amboss.com/us/questions/7B2b04X4Ge/2> | A | A | Infectious Diseases | Correct |
| <https://next.amboss.com/us/questions/7B2b04X4Ge/3> | E | E | Infectious Diseases | Correct |
| <https://next.amboss.com/us/questions/7B2b04X4Ge/4> | A | A | Infectious Diseases | Correct |
| <https://next.amboss.com/us/questions/7B2b04X4Ge/5> | B | A | Infectious Diseases | Incorrect |
| <https://next.amboss.com/us/questions/7B2b04X4Ge/7> | D | B | Infectious Diseases | Incorrect |
| <https://next.amboss.com/us/questions/7B2b04X4Ge/8> | A | A | Infectious Diseases | Correct |
| <https://next.amboss.com/us/questions/7B2b04X4Ge/9> | A | A | Infectious Diseases | Correct |
| <https://next.amboss.com/us/questions/7B2b04X4Ge/10> | A | A | Infectious Diseases | Correct |
| <https://next.amboss.com/us/questions/7B2b04X4Ge/11> | D | D | Infectious Diseases | Correct |
| <https://next.amboss.com/us/questions/7B2b04X4Ge/12> | D | C | Infectious Diseases | Incorrect |
| <https://next.amboss.com/us/questions/HA2b0KO4Ge/1> | A | A | Obstetrics | Correct |
| <https://next.amboss.com/us/questions/HA2b0KO4Ge/2> | D | D | Obstetrics | Correct |
| <https://next.amboss.com/us/questions/S_2b0yn4Ge/1> | F | E | Clinical pathology/Laboratory medicine | Incorrect |
| <https://next.amboss.com/us/questions/S_2b0yn4Ge/2> | B | B | Clinical pathology/Laboratory medicine | Correct |
| <https://next.amboss.com/us/questions/S_2b0yn4Ge/3> | D | D | Clinical pathology/Laboratory medicine | Correct |
| <https://next.amboss.com/us/questions/S_2b0yn4Ge/4> | E | C | Clinical pathology/Laboratory medicine | Incorrect |
| <https://next.amboss.com/us/questions/S_2b0yn4Ge/5> | A | E | Clinical pathology/Laboratory medicine | Incorrect |
| <https://next.amboss.com/us/questions/S_2b0yn4Ge/6> | B | D | Clinical pathology/Laboratory medicine | Incorrect |
| <https://next.amboss.com/us/questions/S_2b0yn4Ge/7> | E | C | Clinical pathology/Laboratory medicine | Incorrect |
| <https://next.amboss.com/us/questions/S_2b0yn4Ge/8> | D | D | Clinical pathology/Laboratory medicine | Correct |
| <https://next.amboss.com/us/questions/S_2b0yn4Ge/9> | B | B | Clinical pathology/Laboratory medicine | Correct |
| <https://next.amboss.com/us/questions/S_2b0yn4Ge/10> | A | A | Clinical pathology/Laboratory medicine | Correct |
| <https://next.amboss.com/us/questions/S_2b0yn4Ge/11> | E | E | Clinical pathology/Laboratory medicine | Correct |
| <https://next.amboss.com/us/questions/S_2b0yn4Ge/12> | F | F | Clinical pathology/Laboratory medicine | Correct |
| <https://next.amboss.com/us/questions/S_2b0yn4Ge/13> | E | E | Clinical pathology/Laboratory medicine | Correct |
| <https://next.amboss.com/us/questions/S_2b0yn4Ge/14> | A | C | Clinical pathology/Laboratory medicine | Incorrect |

### Supplementary Table 2. Adherence Rates to the Criteria in GPT-4V Explanations for Correct and Incorrect AMBOSS Responses.

| Evaluation criteria | Correct response (n=95) | Incorrect response (n=55) |
| --- | --- | --- |
| Image Interpretation | 84.2% | 92.8% |
| Question Information | 96.8% | 89.1% |
| Comprehensive Explanation | 79.0% | 7.2% |

Note: Given each exam question, healthcare professionals evaluate GPT-4V explanations for the following criteria. Image Interpretation: GPT-4V sufficiently supported its choice through its interpretation of the image; Question Information: The explanation provided essential patient information from the textual question context; Comprehensive Explanation: The explanation included comprehensive reasoning for all possible that leads to the final answer.
